# Sensitive periods for prenatal alcohol exposure shape internalizing symptoms across development

**DOI:** 10.64898/2026.06.23.26356366

**Authors:** Ka Ying Toby Law, Madison E. Bigler, Emma Kohrt, Alex S. F. Kwong, Alexandre A. Lussier

## Abstract

**Importance:** Prenatal alcohol exposure (PAE) is associated with lasting cognitive and neurodevelopmental deficits and can quadruple risk for depression later in life. However, it remains unknown whether there are specific trimesters when PAE is more strongly associated with longitudinal trajectories of internalizing symptoms - an indicator of depression risk - across childhood and adolescence.

**Objective:** To investigate how PAE timing and dosage are associated with internalizing symptom trajectories from ages 4 to 16.5 years.

**Design, Setting and Participants:** We analyzed prospective data from the Avon Longitudinal Study of Parents and Children (ALSPAC), an ongoing longitudinal birth cohort from the United Kingdom. Internalizing symptom trajectories were estimated for 6,409 participants. Primary analyses were conducted on 2,254 participants with complete data on PAE in all three trimesters, covariates, and trajectories.

**Main Outcomes and Measures:** We used growth mixture modelling to identify latent trajectories of depressive symptoms measured using the internalizing symptom scale from the Strengths and Difficulties Questionnaire (SDQ) at seven occasions between ages 4 to 16.5 years. Prospective alcohol consumption during each trimester were categorized into three PAE dosages: unexposed (0 drinks/week), low (1-7 drinks/week) and high (7+ drinks/week).

**Results:** We identified five distinct depressive symptom trajectories: stable low (75.9% of participants), moderate childhood peak (11.2%), progressive increase (5.57%), high early childhood (4.73%), and early adolescent peak (2.61%). PAE in the second (relative risk [RR]=2.08, 95% CI=1.15-3.76) and third trimesters (RR=1.83, 95% CI=1.05-3.21), as well as total PAE burden across pregnancy (RR=1.33, 95% CI=1.06-1.68) increased risk for the progressive increase trajectory, versus the stable low trajectory. High PAE in the second (RR=2.71, 95% CI=1.41-5.21) and third (RR=2.27, 95% CI=1.27-4.05) trimesters drove elevated risk for this trajectory. PAE in the first trimester or at low dosages showed no associations with depressive symptom trajectories. Negative control analyses of paternal drinking also found no associations.

**Conclusions and Relevance:** Our results highlight the second and third trimesters as potential sensitive periods for the impact of PAE on rising depressive symptoms from childhood to adolescence. Ultimately, these findings could inform the design of prevention programs, and facilitate targeted interventions to youth at elevated risk for depression.

## INTRODUCTION

Prenatal alcohol exposure (PAE) is associated with a range of cognitive, behavioral, and neurodevelopmental deficits collectively known as Fetal Alcohol Spectrum Disorder (FASD),^1–6^ which is estimated to affect 2-5% of individuals in the USA and 22.8% worldwide.^7–13^ While 20% of people in the general population experience a mental disorder in their lifetime,^14,15^ 90% of individuals with FASD have at least one mental illness in their lifetime,^3–5^ with 50% suffering from depression.^3,4,16–18^ Importantly, mental disorders are also prevalent among people with low levels of PAE, with empirical studies of PAE showing that even moderate levels of PAE can more than double risk for internalizing disorders, such as depression and anxiety.^19–22^ Yet, prior studies investigating the link between PAE and depression have not yet explored two key dimensions of this relationship: 1) the timing of PAE and 2) trajectories of symptoms across development. These gaps limit our ability to identify optimal periods across pregnancy or offspring development that can be leveraged to mitigate the impact of PAE on depression risk.

First, it remains unclear whether there are developmental windows, or sensitive periods, when PAE has greater influences on depressive symptoms across childhood and adolescence. Both animal and human studies have documented potential sensitive periods for the effects of PAE on cognition and behavior.^20,23–35^ Yet, few studies have investigated the time-dependent effects of PAE on depression, with most existing studies relying on single retrospective measures for the entire pregnancy.^5,36–42^ To our knowledge, only one study prospectively examined PAE timing across all three trimesters and depression risk, showing high PAE (7+ drinks/week) in the second and third trimesters is associated with greater offspring internalizing symptoms in early adulthood, compared to no drinking or drinking in the first trimester.^43^ Although this work highlights potential sensitive periods when PAE has stronger influences on adult depression, no studies have examined whether the timing of PAE influences depressive symptoms earlier in development. Importantly, the identification of sensitive periods for PAE could translate to more timely and efficient use of resources to buffer the impact of PAE on mental health.

Second, prior studies of PAE have mainly evaluated offspring mental and behavioral health outcomes at single timepoints.^21,22,43–47^ However, multiple population-level studies have identified distinct trajectories of depressive symptoms across childhood and adolescence. Symptom trajectories often display non-linear patterns throughout development, with symptoms emerging, peaking, and ebbing at different ages.^48–51^ Thus, cross-sectional evaluations of depressive symptoms may miss meaningful differences in onset, progression patterns, and symptom severity across age groups.^20^ Only one study has quantified effects of PAE on internalizing behavior across childhood and adolescence.^52^ Using linear mixed effects modeling, this study showed light/moderate PAE in the first trimester (2-10 drinks/week) associates with fewer total internalizing behaviors between ages 2-14 years, contrasting findings from cross-sectional studies.^47,53^ However, this approach did not reveal when such behavioral differences emerged across development. Thus, it remains unclear when and how depressive symptoms linked to PAE manifest across childhood and adolescence, limiting our ability to identify developmental windows when interventions to treat or prevent depression may be more effective.

To address these gaps, we analyzed data from a large-scale population cohort, the Avon Longitudinal Study of Parents and Children (ALSPAC), to determine when and how PAE influences internalizing symptom trajectories between ages 4-16.5 years. Specifically, we used internalizing symptoms as a proxy for depressive symptoms to capture broader transdiagnostic features of depression in community settings. To our knowledge, this is the first study to integrate both sensitive periods for PAE and trajectories of mental health across childhood and adolescence.

## METHODS

### Study design and participants

We used data from ALSPAC, an ongoing population birth cohort which recruited pregnant mothers resident in Avon, UK with expected delivery dates between April 1991 to December 1992.^54,55^ 14,541 pregnant mothers were enrolled in the study, among which there were 14,062 eligible live births and 13,988 children alive at 1 year of age. Offspring were followed from gestation through adulthood. The study website provides details of all collected and generated data, available through a fully searchable data dictionary and variable search tool https://www.bristol.ac.uk/alspac/researchers/our-data/.

We restricted primary analyses to consenting singleton participants (eligible sample; n=13,646) who had (1) internalizing symptoms measured at least once in both childhood (age 4-9) and adolescence (age 12-16.5) (trajectory construction sample; n=6,409); and (2) PAE reports available for all three trimesters (n=3,422), resulting in an analytic sample size of 2,254 individuals (Appendix 1).

### Procedures

#### Prenatal alcohol exposure (PAE)

Prospective alcohol consumption (average number of drinks/week) was self-reported by mothers three times during pregnancy. We used the gestational week from each report to assign PAE to one of three trimesters (<18 weeks: first trimester; 18-27 weeks: second trimester; >27 weeks: third trimester; Appendix 2, **Figure S1**). Responses collected prior to 18-weeks gestation were categorized as the first trimester to align with the ALSPAC data collection timepoint and maximize report availability, therefore this category should be interpreted as first trimester (0-13 weeks) and early second trimester (14-18 weeks). For each trimester, we collapsed continuous alcohol consumption into categorical PAE levels (Appendix 2, **Figure S2**): Unexposed (0 drinks/week), low (1-7 drinks/week), and high (7+ drinks/week). These thresholds are consistent with no, light/moderate, and heavy alcohol consumption levels that yielded meaningful results in previous studies.^20,21,26,43,45–47,52^

We constructed five main exposure models to test timing-specific associations with PAE: (1-3) sensitive periods for each trimester (ordinal; range 0-2), (4) accumulation (total PAE across pregnancy; ordinal; range 0-6), and (5) ever-exposed (dichotomous PAE; 0/1). To investigate dosage-specific associations, sensitive period models were treated as nominal variables (no, low, or high PAE).

#### Depressive symptoms

Caregivers reported youth internalizing symptoms on seven occasions (4, 7, 8, 9, 12, 13 and 16.5 years old) using the Strength and Difficulties Questionnaire (SDQ). The SDQ is an established screening tool for clinical cases of child mental disorders^47^ that can improve detection of child psychiatric disorders in community settings.^58^ Internalizing symptom scores were derived from the 5-item emotional symptoms and peer problems subscales. Each item is scored from 0 (not true) to 2 (certainly true), summing to a total score ranging from 0 to 20, with higher scores indicating higher symptom levels (Appendix 2). We used the internalizing symptom scale as a proxy for depressive symptoms, as it captures a spectrum of precursory indicators for anxiety-depressive disorders, including emotional distress, excessive rumination, somatic complaints, and social isolation or withdrawal. The internalizing scale also has better construct and discriminant validity for clinical psychiatric disorders in low-risk samples, compared to the emotional symptoms subscale alone.^57,58^

#### Construction of latent internalizing symptom trajectories

We used growth mixture modelling (GMM) in Mplus version 8.11^59^ to construct latent trajectories of internalizing symptoms measured from ages 4 to 16.5 years. Briefly, GMM stratifies heterogenous, individual-level trajectories into homogenous subpopulations,^60^ each represented by a distinct population-level trajectory (or latent class). Participants received a probability of assignment to each derived trajectory class and were given membership to the trajectory with the highest probability. As symptom heterogeneity between individuals can be influenced by social determinants, we used the 1-step approach for GMM, which allows covariates to influence trajectory growth factors and latent class formation. Missingness was addressed using full-information maximum likelihood (FIML) during GMM.^59^ Full details of validity assessments used to determine the best-fit number of latent classes are included in Appendix 3.

#### Covariates

We adjusted for the following covariates in modelling and analyses due to their potential effects on PAE and contributions to internalizing symptom heterogeneity: maternal age at birth, gestational age, child sex, parity, self-reported ethnicity, maternal education at birth, maternal depressive symptoms (Edinburgh Postnatal Depression Scale [EPDS]; 8 weeks postpartum), and smoking during pregnancy (Appendix 2).

### Statistical analyses

#### Primary analyses

We performed multinomial logistic regressions in R (version 4.3.3)^61^ to test the relationship between PAE timing and dosages with internalizing symptom trajectories. Standardized relative risk ratios (RR – exponentiated *log* odds ratio), 95% CI, and P-values were calculated to assess the risk conferred by each PAE model on trajectory assignment, relative to the latent trajectory with the largest class size and no PAE (i.e., reference levels).

#### Sensitivity and negative control analyses

Consistency of findings from the main multinomial logistic regression analyses were validated via the following sensitivity analyses:

i. continuous PAE measures (average drinks/week; Appendix 4);
ii. dichotomously coded sensitive period hypotheses (i.e., no dosage information) and cumulative PAE in the second and third trimesters (Appendix 5); and
iii. the population sample used in trajectory construction (N=6,409; Appendix 6), which includes missing PAE reports.

We also conducted negative control analyses using partner alcohol consumption during pregnancy, as this exposure does not directly impact fetal development (N=6,409; Appendices 2 and 7).

## RESULTS

### Sample description

Of 13,646 eligible participants, 2,254 (49.6% female) had both internalizing symptom trajectories and complete PAE data available for primary analyses (**Table 1**). Compared to the sample used to construct internalizing symptom trajectories, the analytic sample had slightly higher maternal education at birth (Appendix 1). Compared to participants with high PAE, those with no or low PAE tended to have mothers with higher educational attainment and lower EPDS scores at 8 weeks postpartum, and were less likely to be exposed to smoking prenatally (Appendix 8).

**Table 1.**
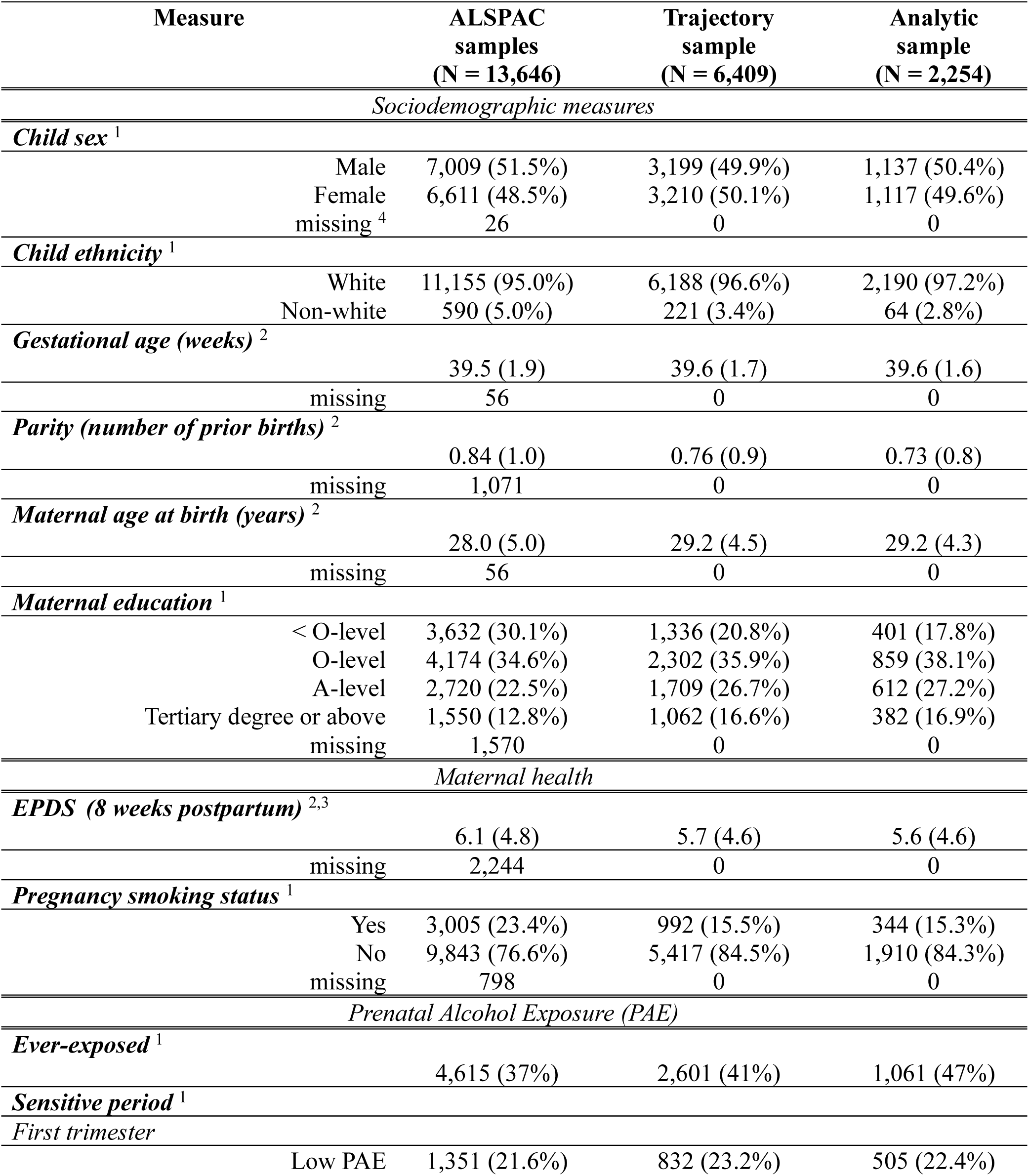
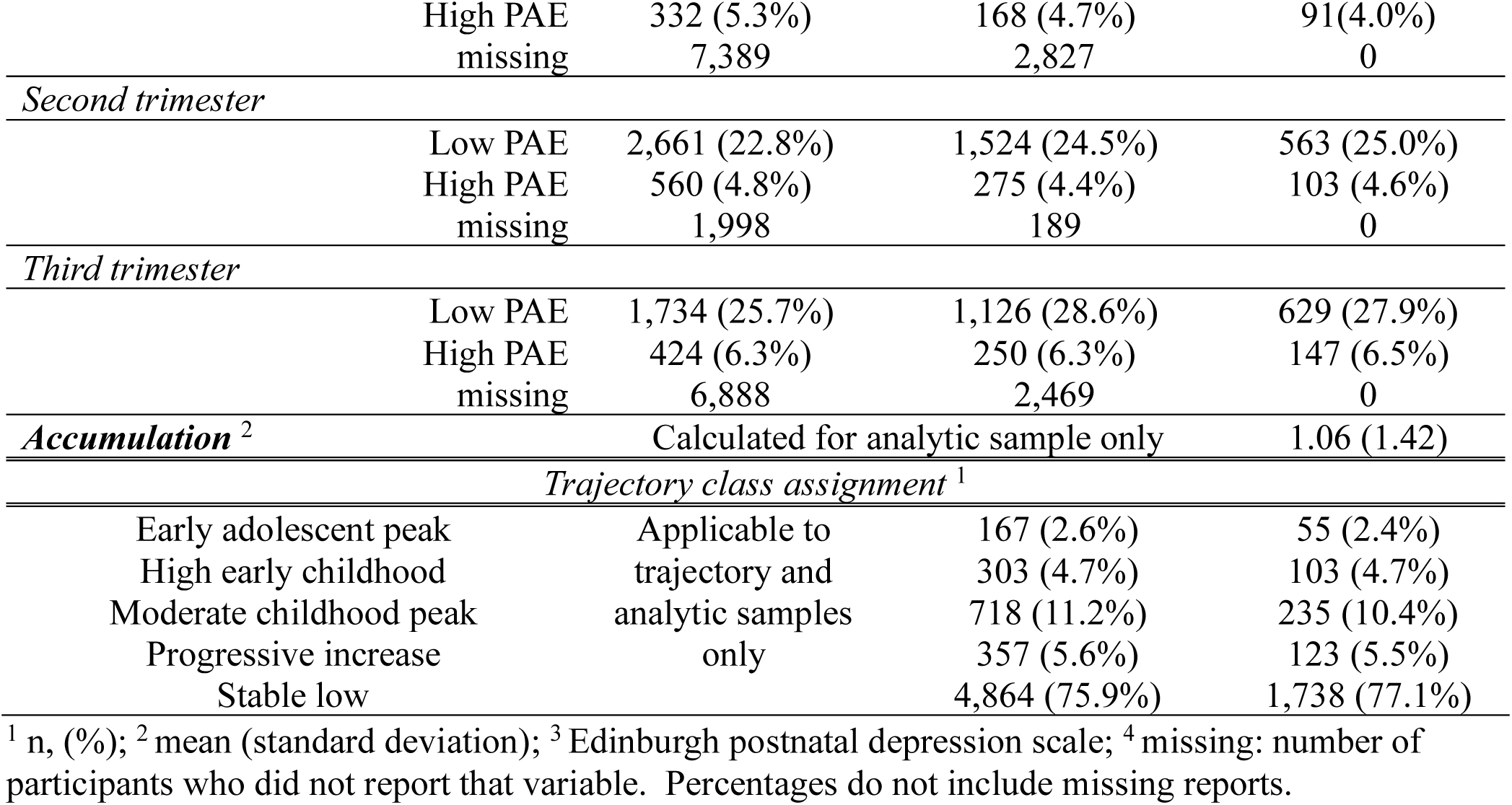
Comparisons between eligible ALSPAC sample, trajectory modelling sample, and analytic sample. Sociodemographic characteristics, maternal health, prevalence of PAE, and trajectory class assignment (trajectory and analytic sample only).

### Internalizing symptom trajectories

Among participants included in trajectory construction, a 5-class solution was the best-fit model of internalizing symptom trajectories (**Figure 1**). Most participants had stable and consistently low symptoms (stable low trajectory, 75.9% of participants) across childhood and adolescence. Two latent trajectories surpassed or approached the validated threshold for possible clinical depression (i.e., ≥9 for caregiver-reported SDQ),^57^ where symptoms peaked in early adolescence (early adolescent peak, 2.61%) or progressively increased (progressive increase, 5.57%) across childhood and adolescent years. The final two trajectories reflected individuals with moderate symptoms peaking in childhood (moderate childhood peak, 11.2%) or higher symptoms in early childhood (high early childhood, 4.73%). Class assignment percentages remained consistent between the trajectory and analytic samples (**Table 1**; **Appendix 1, Table S1**). As the largest latent trajectory class, stable low was set as the reference level for subsequent analyses.

**Figure 1.**
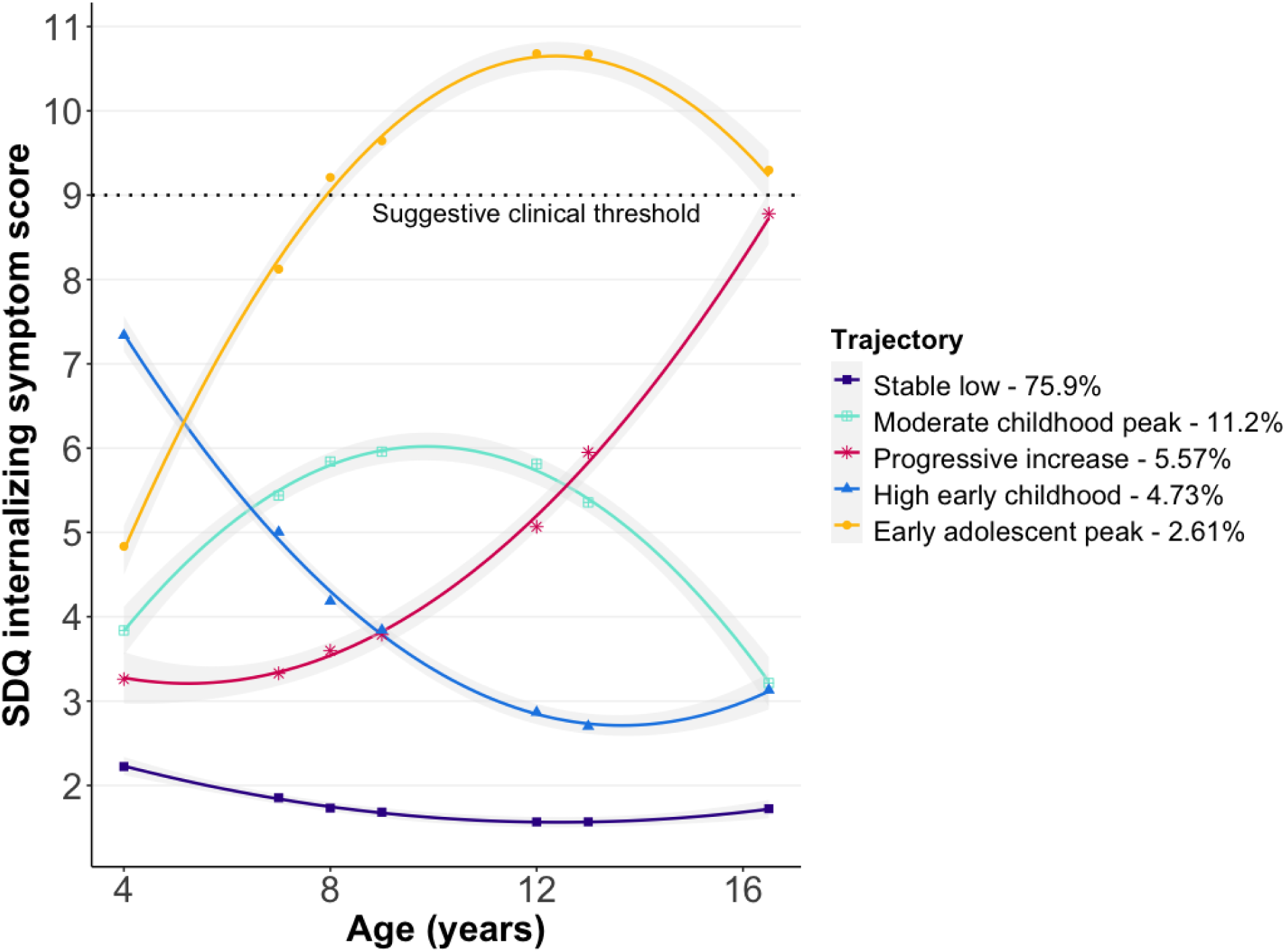
Latent internalizing symptom trajectories from ages 4-16.5 years. Within the trajectory sample of 6,409 individuals, 4,864 (75.9%) were assigned to the stable low trajectory, 718 (11.2%) to the moderate childhood peak trajectory, 357 (5.57%) to the progressive increase trajectory, 303 (4.73%) to the high early childhood trajectory, and 167 (2.61%) to the early adolescent peak trajectory. Internalizing symptom scores from the Strengths and Difficulties Questionnaire (SDQ) were used to index depressive symptoms. 95% confidence intervals of all latent trajectories are shown in grey. Suggestive threshold of caregiver-reported SDQ internalizing symptom scores for clinical depression is ≥9. Class assignment frequencies within the analytic sample of 2,254 individuals are included in Table 1.

### PAE timing and internalizing symptom trajectories

Each stepwise increase in PAE level (from none to low, or low to high) in the second and third trimesters was associated with 2.08 and 1.84 times higher risk for the progressive increase trajectory, respectively (Trimester 2: RR=2.08, p=0.01; Trimester 3: RR=1.84, p=0.03) (**Table 2**; **Figure 2**). Total PAE burden across pregnancy was also associated with 1.33 times higher risk for the progressive increase trajectory per stepwise increase in PAE level (RR=1.33, p=0.02). No associations were observed for PAE in the first trimester, nor for dichotomous PAE across pregnancy (i.e., never/ever-exposed).

**Table 2.** Relative risk ratio (RR) conferred by main PAE models for each internalizing symptom trajectory compared to the stable low class.

| PAE models | Moderate childhood peak | Progressive increase | High early childhood | Early adolescent peak |
| --- | --- | --- | --- | --- |
| *Timing hypotheses* |  |  |  |  |
| *Ever-exposed* | 0.90 [0.51, 1.59]<br>p=0.71 | 1.77 [0.84, 3.73]<br>p=0.13 | 1.60 [-0.71, 3.63]<br>p=0.26 | 0.45 [-1.92, 1.4]<br>p=0.17 |
| *Sensitive period* |  |  |  |  |
| First trimester | 0.72 [0.43, 1.22]<br>p=0.22 | 1.59 [0.85, 2.95]<br>p=0.15 | 0.85 [0.40, 1.81]<br>p=0.68 | 0.55 [0.18, 1.62]<br>p=0.28 |
| Second trimester | 0.96 [0.58, 1.59]<br>p=0.88 | **2.08 [1.15, 3.76] \***<br>**p=0.01** | 1.47 [0.75, 2.90]<br>p=0.26 | 0.90 [0.34, 2.35]<br>p=0.83 |
| Third trimester | 1.01 [0.64, 1.60]<br>p=0.96 | **1.84 [1.05, 3.21] \***<br>**p=0.03** | 1.26 [0.65, 2.41]<br>p=0.49 | 0.91 [0.37, 2.26]<br>p=0.84 |
| *Accumulation* | 0.95 [0.78, 1.16]<br>p=0.62 | **1.33 [1.06, 1.68] \***<br>**p=0.02** | 1.08 [0.82, 1.43]<br>p=0.56 | 0.89 [0.60, 1.32]<br>p=0.56 |
| *Dosage hypotheses* |  |  |  |  |
| *First trimester* |  |  |  |  |
| Low PAE | 0.84 [0.59, 1.20]<br>p=0.34 | 1.29 [0.84, 1.97]<br>p=0.25 | 1.0 [0.61, 1.64]<br>p=1.00 | 0.90 [0.46, 1.74]<br>p=0.75 |
| High PAE | 0.73 [0.36, 1.46]<br>p=0.37 | 1.52 [0.69, 3.35]<br>p=0.30 | 0.71 [0.24, 2.08]<br>p=0.53 | 0.28 [0.04, 2.17]<br>p=0.23 |
| *Second trimester* |  |  |  |  |
| Low PAE | 0.99 [0.71, 1.38]<br>p=0.95 | 1.15 [0.75, 1.77]<br>p=0.53 | 1.29 [0.82, 2.04]<br>p=0.27 | 0.92 [0.48, 1.76]<br>p=0.80 |
| High PAE | 0.95 [0.49, 1.87]<br>p=0.89 | **2.71 [1.41, 5.21] \*\***<br>**p=0.003** | 1.33 [0.54, 3.27]<br>p=0.54 | 0.96 [0.28, 3.31]<br>p=0.94 |
| *Third trimester* |  |  |  |  |
| Low PAE | 0.98 [0.71, 1.36]<br>p=0.92 | 1.02 [0.66, 1.58]<br>p=0.92 | **1.56 [1.00, 2.41] \***<br>**p=0.048** | 0.88 [0.47, 1.68]<br>p=0.71 |
| High PAE | 1.04 [0.60, 1.81]<br>p=0.89 | **2.27 [1.27, 4.05] \*\***<br>**p=0.005** | 0.65 [0.23, 1.86]<br>p=0.42 | 1.01 [0.34, 2.98]<br>p=0.99 |

RR; [95%CI]; p-value. RR: relative risk ratio, derived from exponentiated odds ratio.
\*p<0.05, \*\*p<0.01.

**Figure 2.**
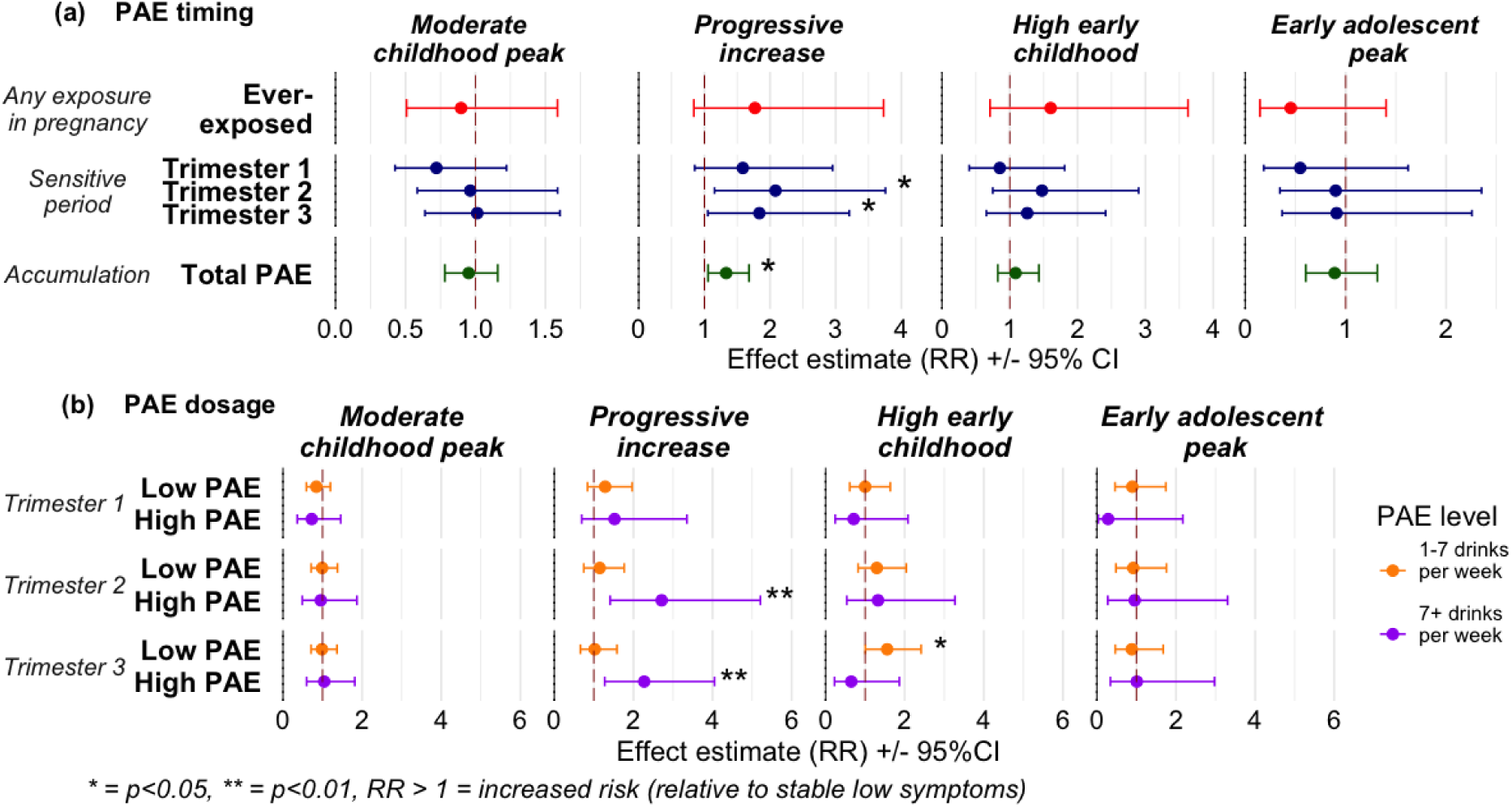
Associations between PAE models and latent internalizing symptom trajectories. Each panel shows the relative risk ratios (RR) and 95% confidence intervals (CI) for each PAE model and internalizing symptom trajectory compared to the reference levels (no PAE, stable low trajectory). **a.** Association with PAE timing models (any exposure in pregnancy, sensitive period exposure, accumulative burden). For sensitive periods, RR reflects risk conferred by each stepwise increase in PAE level (from none to low, or low to high). **b.** Association with PAE dosage models (PAE at low and high dosages in the first, second, and third trimesters of pregnancy), compared to no PAE.

### PAE dosage and internalizing symptom trajectories

Compared to no PAE, only high PAE (7+ drinks/week) in the second and third trimesters was associated with 2.71 and 2.27 times higher risk for the progressive increase trajectory, respectively (Trimester 2: RR=2.71, p=0.003; Trimester 3: RR=2.27, p=0.005). By contrast, low PAE in the third trimester was associated with a marginal risk increase for the high early childhood trajectory (RR=1.56, p=0.048). No associations were observed for either low or high PAE in the first trimester.

### Sensitivity and negative control analyses

Sensitivity analyses using continuous PAE (Appendix 4), cumulative PAE from second and third trimesters (Appendix 5: **Table S5, Figure S4**), and incomplete PAE reports from the trajectory modeling sample (N=6,409; Appendix 6) largely supported the association between second and third trimester PAE with increased relative risk for the progressive increase trajectory. Dichotomous sensitive period exposures were not associated with differences in risk for any trajectory (Appendix 5: **Table S5**, **Figure S5**), highlighting the importance of accounting for PAE dosage. Finally, negative control analyses of partner drinking did not identify any associations with internalizing symptom trajectories, regardless of timing or level, reducing potential concerns of environmental confounding (Appendix 7).

## DISCUSSION

In this study, we examined the relationship between PAE timing and dosage and depressive symptom trajectories between ages 4-16.5 years. Specifically, we identified the second and third trimesters of pregnancy as potential sensitive periods when higher PAE levels are associated with greater risk of progressively increasing internalizing symptoms across childhood and adolescence.

To our knowledge, we are the first to demonstrate that the timing of PAE is associated with longitudinal depressive symptom emergence, progression, and severity. Our discovery of sensitive periods in trimesters two and three aligns with previous evidence that the consequences of PAE vary by gestational timing,^20,23–34^ likely because neurobiological systems undergo distinct stages of development during different periods in gestation.^62,63^ Lack of associations with partner drinking further supports that timing effects are specific to biological exposure *in utero* and not parent behavioral or environmental confounding. The second and third trimesters of human pregnancy are peak periods for synaptogenesis and regional brain connectivity, and the third trimester involves rapid neuronal proliferation and gyrification.^64,65^ Lower brain volume, abnormal cortical structures and gyral patterns, and altered vascular organization are observed in people with PAE and FASD.^66–68^ Similarly, patients with major depressive disorder display altered regional grey-matter volumes and neuronal signaling synchrony, as well as abnormal cerebral blood perfusion.^69,70^ Thus, PAE in the second and third trimesters may produce persistent deficits in brain connectivity, vascularization, and morphology relevant to pathophysiology of both PAE and depression, which may explain the time-specific association between PAE and rising depressive symptoms. By contrast, we did not detect any associations with first trimester PAE, which is generally associated with physical symptoms, such as craniofacial, skeletal, and internal organ dysmorphology.^27,35^ Therefore, PAE limited to early pregnancy may be less relevant for behavioral and neurological sequelae, suggesting screening for mental disorders following PAE should focus on youth exposed during the second and/or third trimesters.

The dose-dependent impact of PAE on risk for psychiatric diagnoses is well documented, regardless of trimester and prospective^21,22,43,47,52^ or retrospective^36,37,39–41,71^ measures. In addition to confirming PAE dose-dependency within a population-based cohort, we further revealed that sensitive period exposure *in tandem* with high dosage was most strongly associated with rising internalizing symptoms across development. Although we did not identify associations with low PAE, our findings do not necessarily support a safe PAE threshold, as higher socioeconomic position (SEP) – a predictor of positive child outcomes – was confounded with low PAE in our study (Appendix 8), a common pattern across multiple studies and cohorts.^39,52,72^ Given that offspring mental health is heavily influenced by social and rearing environments,^73–75^ smaller negative effects of low PAE can be masked or mitigated by protective effects of high SEP.^39,45^ Thus, absence of population-level associations with low PAE do not rule out that some individuals may still be susceptible to its impacts.^76^

Our focus on depressive symptom trajectories allowed us to characterize how PAE shapes the magnitude, timing, and progression of symptoms throughout development. This approach is of high clinical value, as it demonstrates how youth with elevated depression risk may present symptomatically over time and reveal optimal windows for intervention. From the shape of the progressive increase trajectory, it appears the time-dependent effects of PAE on internalizing symptoms are latent, rather than immediate – after initial exposure, symptom levels remained in the normal range through childhood, then increased substantially upon reaching adolescence. These results could be explained by the sensitization of children and adolescents with PAE to environmental insults, which are further compounded by stressors of pubertal development. PAE can delay neurobiological mechanisms governing elevated neuroplasticity, which could stunt learning, resilience-building and adaptation to the postnatal environment.^77^ PAE is also associated with hyperresponsiveness and dysregulation of the stress response,^14^ altering the ability to process and respond to acute and chronic environmental stressors,^78^ which may increase susceptibility for internalizing disorders. In parallel, the pubertal period presents a series of novel challenges such as changes in physique, hormonal and mood shifts, academic pressures, changing family and peer relationships.^79^ Beyond these developmental shifts, youth with PAE also often face disproportionately higher levels of adverse childhood experiences (ACEs), which could further explain the association between PAE and progressively rising depressive symptoms.^80^ Therefore, heightened sensitivity to stress and adversity, combined with typical social and developmental challenges, might explain the relationship between PAE and a gradual rise in depressive symptoms across development.

From a clinical perspective, the findings of this study reveal valuable windows of opportunity where interventions or screening programs could effectively mitigate the impact of PAE on youth depression risk. For example, prospective screening for high PAE during sensitive periods in pregnancy could help to identify expectant parents most in need of social support programs and medical guidance surrounding alcohol misuse or reliance. PAE can also interact with ACEs to increase vulnerability to depression, anxiety, and psychotic symptoms.^81,82^ Retrospective screening for second or third trimester PAE, alongside a history of ACEs could also help identify and target interventions to children and adolescents who are most at risk for developing clinical depression, particularly among those without an FASD diagnosis.

## LIMITATIONS

Our study had some limitations. First, ALSPAC is primarily a white European cohort with relatively high SEP, and missingness in PAE reports was partially related to lower SEP during gestation (**Table 1**; Appendix 1, **Table S1**). Thus, our results should be replicated in more diverse populations to ensure their generalizability across different sociodemographic contexts. Second, analytic samples were restricted by lower response rates for both PAE and youth internalizing symptoms (**Table 1**), such that only 2,254 out of the 6,409 participants included in trajectory modelling remained for association analyses. This difference could be attributed to participant attrition over time^83^ and stigma surrounding PAE and/or mental health. Nonetheless, we found largely consistent class assignment frequencies (Table 1) and associations with PAE timing and dosage across the analytic and trajectory modelling samples (Appendix 6). Third, although we conducted the largest analysis of repeated, prospective PAE and longitudinal mental health within the same participants, we could not fully evaluate the impact of PAE in the context of other determinants of depression. For instance, not all latent trajectories of interest maintained sufficient class frequencies to conduct interaction analyses with child sex or sex-stratified analyses, despite known sex differences in depression.^39,51,84^ Lastly, we used the most probable trajectory class assignments as our outcome variable, which may omit class uncertainty and incorporate a larger error in our results. However, average modal posterior probabilities for all five latent classes were above 75% (range=75.2–94.2%; Appendix 9), indicative of good prediction certainty and limiting this potential concern.^60^

## CONCLUSIONS

Our findings suggest PAE has nuanced and time-specific effects on depressive symptom development, progression, and severity across childhood and adolescence, with trimesters two and three identified as potential sensitive periods when high PAE has greater impacts on internalizing symptom trajectories. Ultimately, these sensitive periods for PAE and their association with depression risk could help establish screening programs that identify and deliver timely interventions to expecting families, as well as youth at elevated risk for depression.

## DISCLOSURES

The authors have no conflicts of interest to declare.

## ACKNOWLEDGEMENTS

This work was supported by the National Institute on Alcohol Abuse and Alcoholism (NIAAA) of the National Institutes of Health (grant number R21AA030640 awarded to AAL). The content is solely the responsibility of the authors and does not necessarily represent the official views of the National Institutes of Health. Dr. Lussier is supported by an MQ Fellows Award from the MQ Foundation (MQF22\9). Dr. Kwong is supported by a Wellcome Early Career Award (Grant ref: 227063/Z/23/Z). We thank Dr. Andrew D.A.C Smith and Dr. Joanne Weinberg for their input and assistance with the conceptualization of PAE variables.

We are extremely grateful to all the families who took part in this study, the midwives for their help in recruiting them, and the whole ALSPAC team, which includes data collection staff, data and administrations staff, technical managers and the technical staff with the Bristol Bioresource Laboratory, based within the University of Bristol. The UK Medical Research Council and Wellcome (Grant ref: MR/Z505924/1) and the University of Bristol provide core support for ALSPAC. This publication is the work of the authors, who will serve as guarantors for the contents of this paper.

## DATA AVAILABILITY

The informed consent obtained from ALSPAC participants does not allow the data to be made available through any third party maintained public repository. All data are available by request from the ALSPAC Executive Committee for researchers who meet the criteria for access to confidential data (https://bristol.ac.uk/alspac/researchers/access). The ALSPAC study website contains details of all available data (https://www.bristol.ac.uk/alspac/researchers/our-data/).

## INCLUSION AND ETHICS

Ethical approval for the study was obtained from the ALSPAC Ethics and Law Committee and the Local Research Ethics Committees. Informed consent for the use of data collected via questionnaires and clinics was obtained from participants following the recommendations of the ALSPAC Ethics and Law Committee. Study participation was voluntary, and participants provided informed consent for the intended use of all data collected during collection sweeps. The completion of a questionnaire, either on paper or online, was considered to be written consent from participants to use their data for research purposes. Data in this study were fully anonymized, and participants can contact the study team at any time to retrospectively withdraw consent for their data to be used. Secondary analyses of these data were approved with oversight by the Mass General Brigham Institutional Review Boards (Protocol 2023P000011).

## Notes

### Competing Interest Statement

The authors have declared no competing interest.

### Author Declarations

Data in this study were fully anonymized, and participants can contact the study team at any time to retrospectively withdraw consent for their data to be used. Secondary analyses of these data were approved with oversight by the Mass General Brigham Institutional Review Boards (Protocol 2023P000011).

