## Supplemental material for "Sensitive periods for prenatal alcohol exposure shape internalizing symptoms across development"

**Appendix 1: Participant differences in trajectory construction and analytic samples.**

We conducted Pearson's chi-squared and Wilcoxon rank-sum tests to summarize participant differences in the trajectory construction and analytic sample. This analysis provided insight on what was driving missingness in PAE reports, and confirmed generalizability of study results to the trajectory construction sample.

**Table S1. Comparisons between trajectory construction sample and analytic sample participants.** Sociodemographic characteristics, maternal health, prevalence of PAE, and trajectory class assignments.

| Measure | Trajectory sample (N = 6,409) | Analytic sample (N = 2,254) | p-value [3] |
|---|---|---|---|
| *Sociodemographic measures* | | | |
| ***Child sex*** [1] | | | 0.7 |
| Male | 3,199 (49.9%) | 1,137 (50.4%) | |
| Female | 3,210 (50.1%) | 1,117 (49.6%) | |
| missing[5] | 0 | 0 | |
| ***Child ethnicity*** [1] | | | 0.2 |
| White | 6,188 (96.6%) | 2,190 (97.2%) | |
| Non-white | 221 (3.4%) | 64 (2.8%) | |
| ***Gestational age (weeks)*** [2] | 39.6 (1.7) | 39.6 (1.6) | 0.2 |
| missing | 0 | 0 | |
| ***Parity (number of prior births)*** [2] | 0.76 (0.85) | 0.73 (0.82) | 0.3 |
| missing | 0 | 0 | |
| ***Maternal age at birth (years)*** [2] | 29.2 (4.5) | 29.2 (4.3) | 0.7 |
| missing | 0 | 0 | |
| ***Maternal education*** [1] | | | 0.017 |
| < O-level | 1,336 (20.8%) | 401 (17.8%) | |
| O-level | 2,302 (35.9%) | 859 (38.1%) | |
| A-level | 1,709 (26.7%) | 612 (27.2%) | |
| Tertiary degree or above | 1,062 (16.6%) | 382 (16.9%) | |
| missing | 0 | 0 | |
| *Maternal health* | | | |
| ***EPDS*** [4] ***(8 weeks postpartum)*** [2] | 5.7 (4.6) | 5.6 (4.6) | 0.054 |
| missing | 0 | 0 | |
| ***Pregnancy smoking status*** [1] | | | 0.8 |
| Yes | 992 (15.5%) | 344 (15.3%) | |
| No | 5,417 (84.5%) | 1,910 (84.7%) | |
| missing | 0 | 0 | |
| *PAE* | | | |
| ***Ever-exposed*** [1] | | | <0.001 |

| | | | |
|---|---|---|---|
| | 2,601 (40.6%) | 1,061 (47.1%) | |
| ***Sensitive period*** [1] | | | |
| *First trimester* | | | 0.3 |
| Low PAE | 832 (23.3%) | 505 (22.4%) | |
| High PAE | 168 (4.7%) | 91 (4.0%) | |
| missing | 2,827 | 0 | |
| *Second trimester* | | | 0.8 |
| Low PAE | 1,524 (24.5%) | 563 (25.0%) | |
| High PAE | 275 (4.4%) | 103 (4.6%) | |
| missing | 189 | 0 | |
| *Third trimester* | | | 0.8 |
| Low PAE | 1,126 (28.6%) | 629 (27.9%) | |
| High PAE | 250 (6.3%) | 147 (6.5%) | |
| missing | 2,469 | 0 | |
| ***Accumulation*** [2] | | | / |
| | Calculated for analytic sample only | 1.06 (1.42) | |
| | *Trajectory class assignment* [1] | | |
| Early adolescent peak | 167 (2.6%) | 55 (2.4%) | 0.8 |
| High early childhood | 303 (4.7%) | 103 (4.7%) | |
| Moderate childhood peak | 718 (11.2%) | 235 (10.4%) | |
| Progressive increase | 357 (5.6%) | 123 (5.5%) | |
| Stable low | 4,864 (75.9%) | 1,738 (77.1%) | |

[1] n, (%); [2] mean (standard deviation); [3] Wilcoxon rank sum test (continuous), Pearson's chi-squared test (categorical); [4] Edinburgh postnatal depression scale; [5] missing: number of participants who did not report that variable. Percentages do not account for missing reports.

**Appendix 2: Supplemental methods for PAE, depressive symptoms, covariates and partner drinking measures.**

*PAE: Timepoint-recoding and categorization*

Prospective alcohol consumption levels were reported by mothers via questionnaires at study intake, and repeated in each subsequent trimester of pregnancy until birth. Participants were advised that one drink was equivalent to 10 grams of pure alcohol, a half-pint of beer/lager, one glass of wine, and one pub-measure of spirits or other alcoholic drinks. Responses were collected in ALSPAC questionnaires A, B, and C, each wave containing reports from a mixture of gestational stages after conception (Figure S1). We used week of gestation at questionnaire completion (ALSPAC variables a902, b924, c991) to assign prospective alcohol consumption reports to pregnancy trimesters (<18 weeks: first trimester; 18-27 weeks: second trimester; >27 weeks: third trimester). Reports prior to 18 weeks gestation were treated as from the first trimester in the ALSPAC study, instead of the conventional clinical cutoff at 13 weeks. Postpartum responses were treated as retrospective reports and excluded from analyses.

Distribution of continuous PAE reports were zero-skewed (i.e. most children were not exposed to alcohol during gestation), which was partially normalized by discretization, improving the representation of observations with PAE (Figure S2).

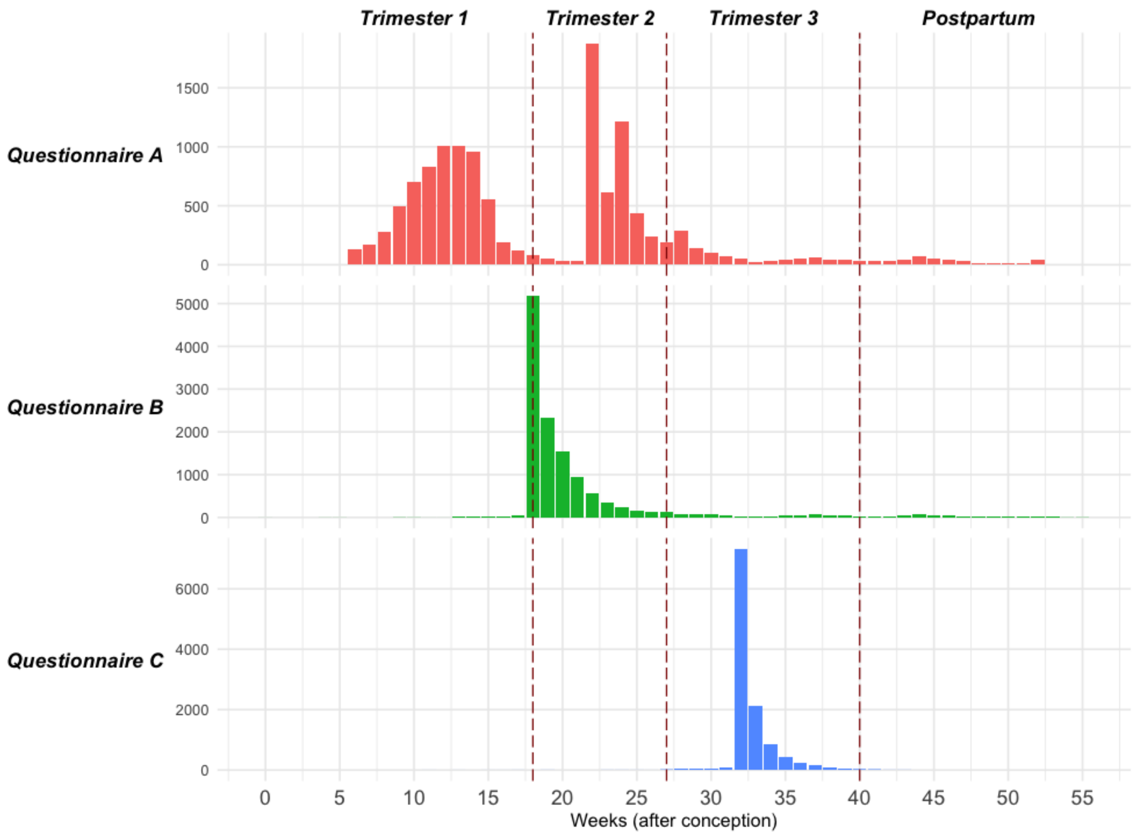

**Figure S1:** Distribution of participant gestational stages in ALSPAC data collection questionnaires A, B, and C, all eligible samples (N=13,646).

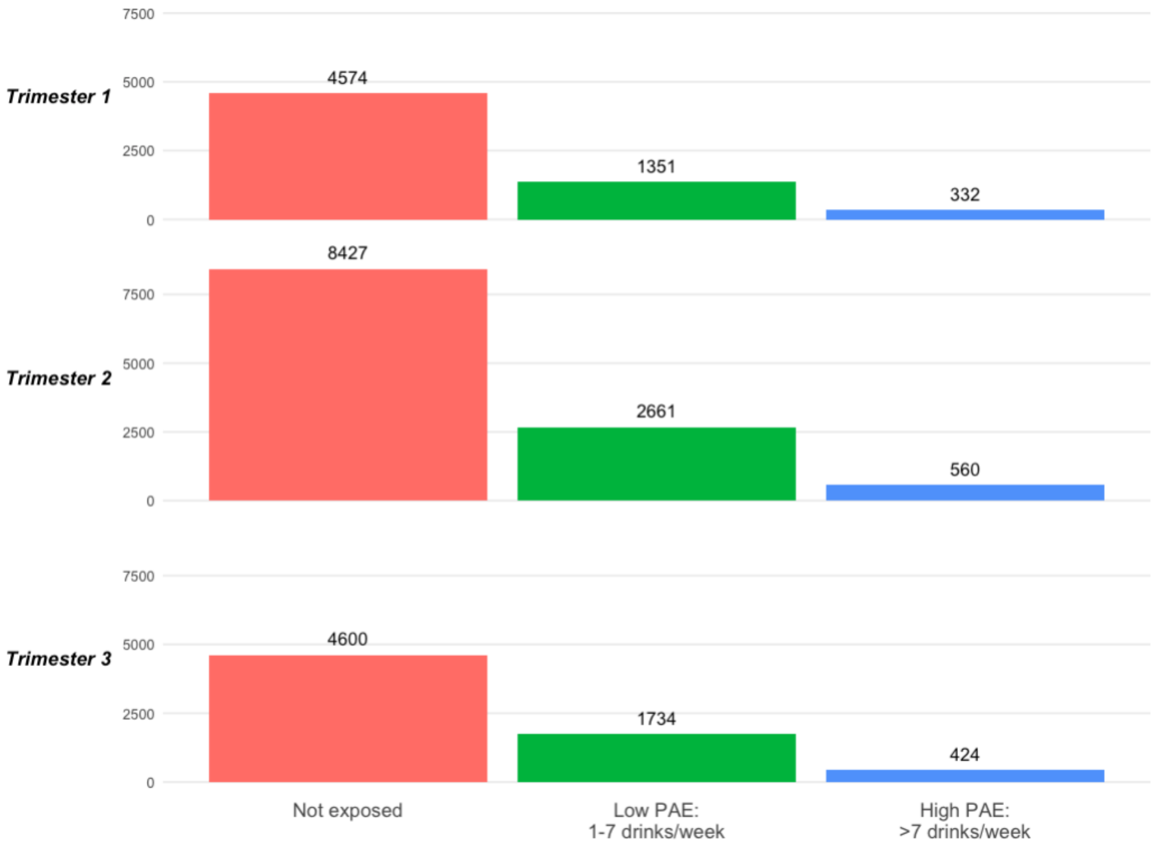

**Figure S2:** Number of participants across categorical PAE levels (not exposed, low PAE, high PAE) after timepoint-recoding and discretization, all eligible samples (N=13,646).

*Depressive symptoms*

Prorated internalizing symptom scores were calculated for participants who partially completed the SDQ emotional symptoms and peer problems subscales. Observations missing more than two items in either subscale were excluded from analysis.

*Covariates*

Mothers' educational attainment and parity were self-reported during study intake. Categories with low frequencies were collapsed into broader classes to reduce model complexity and aid GMM convergence. For educational attainment, the Certificate of Secondary Education (CSE)/no educational qualification class (20.7% of responses) was collapsed with the vocational qualification class (9.7%). Higher educational attainment classes include O-level (34.1%), A-level (22.1%) and tertiary degree (12.7%). O-levels are roughly equivalent to completing middle school (grades 9-10) in the US education system, and A-levels are equivalent to a US high school diploma (grades 11-12).

For parity, reports ranged from 0 to 22, therefore mothers with three or more previous pregnancies (approx. 6%) were combined into the same class.

Child race/ethnicity was derived by ALSPAC from the parents' race/ethnicity backgrounds. Due to the low prevalence of non-white participants, race/ethnicity was coded as white or non-white.

Child biological sex was identified from birth notifications taken around time of delivery, and coded as male or female.

Maternal depressive symptoms were measured using the Edinburgh Postnatal Depression Scale administered via questionnaires at eight weeks postpartum. The 30-point scale consists of 10 items each scored from 0 to 3, with higher scores indicating higher depressive symptoms.

Prospective smoking status and/or smoking frequency during pregnancy were reported by mothers during ALSPAC questionnaires A, B, C, in a similar manner to alcohol consumption levels. Therefore, we adopted the same timepoint recoding approach to assign smoking reports to pregnancy trimesters. Smoking status during pregnancy was coded as a dichotomous variable (0/1) – set as true if mothers' reported smoking in one or more trimesters, and false otherwise.

*Constructing timing and level models from partner drinking reports for negative control analyses*

Prospective alcohol consumption levels were reported by mothers' partners via questionnaires during pregnancy and soon after birth. Responses were collected in ALSPAC questionnaire PB and PC, each containing reports from a mixture of gestational stages after conception in the same manner as maternally-reported alcohol consumption. Therefore, we applied the same timepoint-recoding and categorization procedures as with PAE reports, to create partner drinking models analogous to the PAE models. To maximize the availability of responses for the third trimester, we used retrospective alcohol consumption reports obtained up to 12 weeks postpartum, and the rest were excluded from analyses.

For each trimester, we collapsed continuous alcohol consumption into categorical drinking levels: No (0 drinks/week), low (1-7 drinks/week), and high drinking (7+ drinks/week). As there were insufficient reports available in the first trimester, and none of the participants had complete partner drinking reports across all three trimesters, we were only able to test for timing-specific effects with sensitive period models for the second and third trimesters. These two models were also treated as nominal variables (no, light or heavy drinking) to investigate effects of drinking levels.

To make use of all available partner drinking reports while maintaining comparability with the PAE analyses, we conducted regression analyses within the trajectory construction sample (N=6,409).

**Appendix 3: GMM supplemental methods and results for 1- to 6-class solutions.**

*Growth Mixture Modelling*

Trajectories of depressive symptoms were estimated from SDQ internalizing symptom scores collected at seven timepoints across ages 4 to 16.5 years (Table S2). Intercept, slope and quadratic growth factors were estimated for each class to allow for non-linear trajectory progressions. To reduce convergence issues, growth factor variances are constrained to be equal across trajectories, and variance of the quadratic growth factor within trajectories is set to 0.[1,2]

Starting from a single trajectory ($k$ = 1-class), we added trajectories into the model in a stepwise manner, until best-fit number of trajectories was reached. To assess the optimum, we used the adjusted likelihood ratio test (LRT) proposed by Vuong-Lo-Mendell-Rubin (VLMR) for hypothesis testing, which compares model fit for $k$ versus $k$-1 trajectories. An LRT $p$-value of less than 0.05 suggests a significant improvement in model fit for $k$ over $k$-1 trajectories, and this is repeated until the VLMR-LRT results in non-significance, indicating that the $k$-1 class model is the best-fit number of latent trajectories within our sample (Table S3). We also considered for lowest sample size adjusted Bayesian Information Criterion (ssaBIC), significant Bootstrapped Likelihood Ratio Test (BLRT) statistics, and high entropy values (approaching 1.0, indicating better separation between latent classes).

After determining the optimal number of trajectories, covariates were added into the best-fit model. Since heterogeneity in depressive symptoms could be driven by sociodemographic differences as well as social determinants of health (captured by our list of covariates), we elected for the 1-step approach for GMM which allowed covariates to directly influence trajectory formation and stratification into latent classes alongside internalizing symptoms.[5]

The 4- and 5-class solutions had comparable entropy and BLRT p-values, with the ssaBIC and VLMR-LRT p-values slightly favoring 5-class for model-fit and parsimony. Although the smallest class (corresponding to the early adolescent peak trajectory) had <5% prevalence, this is consistent with lower detection and diagnostic rates of clinical depression in pre-pubescent and adolescent demographics compared to adults,[6–10] which has also been observed in ALSPAC.[11] Within our data, prevalence of SDQ internalizing symptoms scores above the caregiver-reported threshold for possible clinical depression (Table S2), as well as the class prevalence of the highest depressive symptom trajectory in 3-, 4-, and 5- class solutions, are all consistently <5% (Table S3). All of the above theoretically support the early adolescent peak trajectory class to be a legitimate subpopulation in a younger age demographic, rather than an artifact of over-extraction.

**Table S2: Descriptive statistics of SDQ internalizing symptom scores in the eligible ALSPAC sample, trajectory modelling sample and analytic sample.**

| Timepoint | ALSPAC sample (N = 13,646) | Trajectory sample (N = 6,409) | Analytic sample (N = 2,254) |
|---|---|---|---|
| *Age 4* | | | |
| n | 9,252 | 6,002 | 2,160 |
| Mean (SD) | 2.99 (2.41) | 2.88 (2.35) | 2.76 |
| % ≥9 | 3.0% | 2.7% | 3.0% |
| *Age 7* | | | |
| n | 8,205 | 5,801 | 2,072 |
| Mean (SD) | 2.57 (2.55) | 2.50 (2.48) | 2.38 (2.43) |
| % ≥9 | 3.3% | 3.0% | 2.4% |
| *Age 8* | | | |
| n | 7,375 | 5,501 | 1,986 |
| Mean (SD) | 2.97 (2.78) | 2.88 (2.74) | 2.75 (2.62) |
| % ≥9 | 4.7% | 4.4% | 3.9% |
| *Age 9* | | | |
| n | 7,542 | 5,712 | 2,035 |
| Mean (SD) | 2.64 (2.73) | 2.54 (2.64) | 2.40 (2.53) |
| % ≥9 | 4.5% | 4.0% | 3.6% |
| *Age 12* | | | |
| n | 6,872 | 5,704 | 2,043 |
| Mean (SD) | 2.58 (2.74) | 2.50 (2.69) | 2.39 (2.69) |
| % ≥9 | 4.5% | 4.1% | 4.2% |
| *Age 13* | | | |
| n | 6,601 | 5,487 | 1,963 |
| Mean (SD) | 2.64 (2.79) | 2.61 (2.78) | 2.56 (2.79) |
| % ≥9 | 4.7% | 4.6% | 4.3% |
| *Age 16.5* | | | |
| n | 5,310 | 4,499 | 1,649 |
| Mean (SD) | 2.59 (2.78) | 2.53 (2.72) | 2.48 (2.70) |
| % ≥9 | 4.3% | 4.0% | 4.2% |

n: number of participants reporting SDQ the timepoint; SD: standard deviation; % ≥9: percentage of participants with internalizing symptom scores above the validated caregiver-reported threshold for possible clinical depression.

**Table S3: Growth mixture modelling of SDQ internalizing symptoms - Results for 1- to 6-class solutions.**

| *k* | Smallest class % | Number of free parameters | Entropy | ssaBIC | VLMR-LRT (*p*-value) | BLRT p-value |
|---|---|---|---|---|---|---|
| 1 | - | 7 | - | 201039.6 | - | - |
| 2 | 8.3% | 11 | 0.885 | 198401.3 | <.0001 | <.001 |
| 3 | 4.5% | 15 | 0.894 | 197374.0 | <.0001 | <.001 |
| 4 | 3.0% | 19 | 0.857 | 196722.7 | 0.0003 | <.001 |
| **5** | **2.2%** | **23** | **0.853** | **166130.9** | **0.0282** | **<.001** |
| 6 | 2.2% | 27 | 0.835 | 196091.2 | 0.1810 | <.001 |

k: number of latent classes; ssaBIC: sample size adjusted Bayesian Information Criteria; VLMR-LRT: Vuong-Lo-Mendell-Rubin likelihood ratio test.; BLRT: Bootstrapped Likelihood Ratio Test.

**Appendix 4: Sensitivity analysis with continuous PAE within the analytic sample (N=2,254).**

To ensure that categorization of PAE measures into discrete levels of no, low, and high PAE did not discard meaningful associations with depressive symptom trajectories, or arbitrarily assume greater similarity within versus between PAE levels, we conducted regression analyses using continuous PAE measures (average number of drinks per week) in the sensitive period and accumulation models.

**Table S4**: **Relative risk ratio (RR) conferred by continuous PAE measures for each depressive symptom trajectory compared to the stable low class.** Continuous PAE measures (average number of drinks per week) were used to construct sensitive period and accumulation models.

| PAE models | Moderate childhood peak | Progressive increase | High early childhood | Early adolescent peak |
|---|---|---|---|---|
| ***Ever-exposed*** | | | | |
| | 0.95 [0.71, 1.26] p=0.71 | 1.33 [0.92, 1.93] p=0.13 | 1.27 [0.84, 1.90] p=0.26 | 0.67 [0.38, 1.18] p=0.17 |
| ***Sensitive period*** | | | | |
| First trimester | 0.94 [0.87, 1.03] p=0.19 | 1.02 [0.93, 1.13] p=0.63 | 0.99 [0.90, 1.09] p=0.86 | 0.91 [0.75, 1.10] p=0.34 |
| Second trimester | 0.95 [0.87, 1.05] p=0.33 | 1.07 [0.99, 1.16] p=0.07 | 0.99 [0.87, 1.12] p=0.84 | 0.94 [0.79, 1.13] p=0.53 |
| Third trimester | 0.98 [0.90, 1.06] p=0.59 | **1.12 [1.03, 1.22] ** p=0.007** | 0.99 [0.87, 1.11] p=0.82 | 0.96 [0.82, 1.13] p=0.66 |
| ***Accumulation*** | | | | |
| Pregnancy | 0.98 [0.95, 1.01] p=0.26 | **1.04 [1.00, 1.08] * p=0.038** | 0.99 [0.95, 1.04] p=0.81 | 0.97 [0.91, 1.04] p=0.42 |
| Second + third trimester | 0.98 [0.93, 1.03] p=0.41 | **1.06 [1.01, 1.11] ** p=0.009** | 0.99 [0.93, 1.06] p=0.80 | 0.97 [0.89, 1.07] p=0.55 |

RR; [95%CI]; p-value. RR: relative risk ratio, derived from exponentiated log odds ratio.

*p<0.05, **p<0.01.

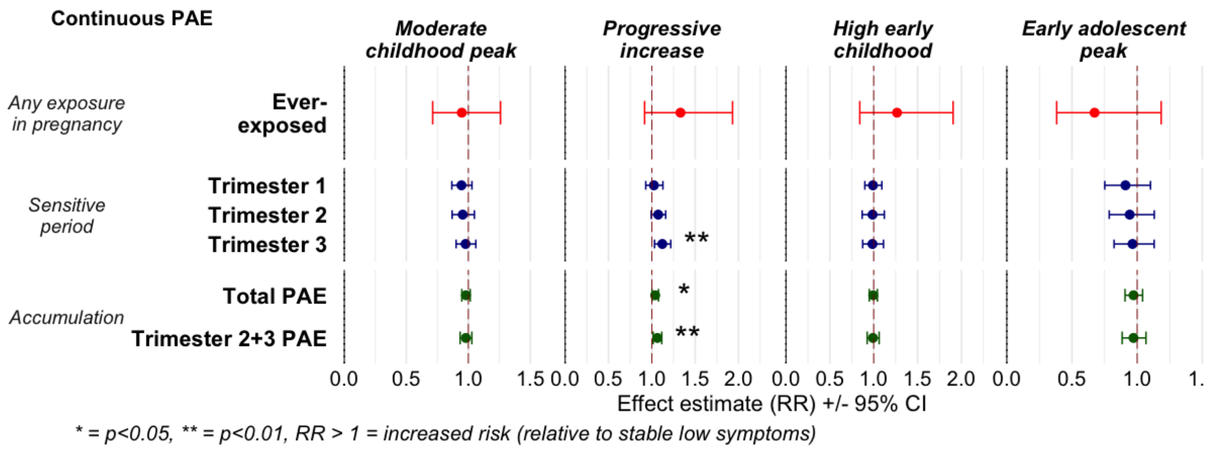

**Figure S3: Associations between continuous PAE models and latent depressive symptom trajectories (relative risk ratios RR, 95% CI).** Continuous PAE measures (average number of drinks per week) were used to construct sensitive period and accumulation hypotheses. For sensitive period and accumulation models, RR reflects risk conferred by each additional drink per week. Top to bottom: associations with any exposure in pregnancy, sensitive periods, accumulative burden across pregnancy or in the second and third trimesters.

**Appendix 5: Sensitivity analyses with alternative PAE models within the analytic sample (N=2,254).**

To validate significant findings between main PAE models and depressive symptom trajectories, we constructed four alternative PAE models to evaluate (1) if timing alone (i.e., PAE without dosage information) was associated with depressive symptom trajectories, (2) how risk conferred by accumulative PAE burden in the second and third trimesters compared to those by other PAE models.

**Table S5**: **Relative risk ratio (RR) conferred by alternative PAE models for each depressive symptom trajectory compared to the stable low class.** Alternative PAE models include dichotomous sensitive period exposure models, and accumulative PAE burden in the second and third trimesters.

| **PAE models** | **Moderate childhood peak** | **Progressive increase** | **High early childhood** | **Early adolescent peak** |
|---|---|---|---|---|
| ***Sensitive period exposure (dichotomous)*** | | | | |
| First trimester | 0.68 [0.35, 1.30] p=0.24 | 1.76 [0.79, 3.89] p=0.16 | 0.90 [0.35, 2.29] p=0.83 | 0.61 [0.17, 2.21] p=0.45 |
| Second trimester | 0.96 [0.52, 1.79] p=0.90 | 1.88 [0.87, 4.09] p=0.11 | 1.69 [0.71, 3.98] p=0.23 | 0.85 [0.26, 2.86] p=0.80 |
| Third trimester | 0.99 [0.54, 1.81] p=0.97 | 1.60 [0.74, 3.45] p=0.23 | 1.88 [0.81, 4.36] p=0.15 | 0.83 [0.25, 2.68] p=0.75 |
| ***Accumulation*** | | | | |
| Second + third trimester | 0.99 [0.76, 1.30] p=0.97 | **1.51 [1.10, 2.08] * p=0.011** | 1.21 [0.83, 1.76] p=0.31 | 0.94 [0.56, 1.58] p=0.82 |

RR; [95%CI]; p-value. RR: relative risk ratio, derived from exponentiated log odds ratio.

*p<0.05, **p<0.01.

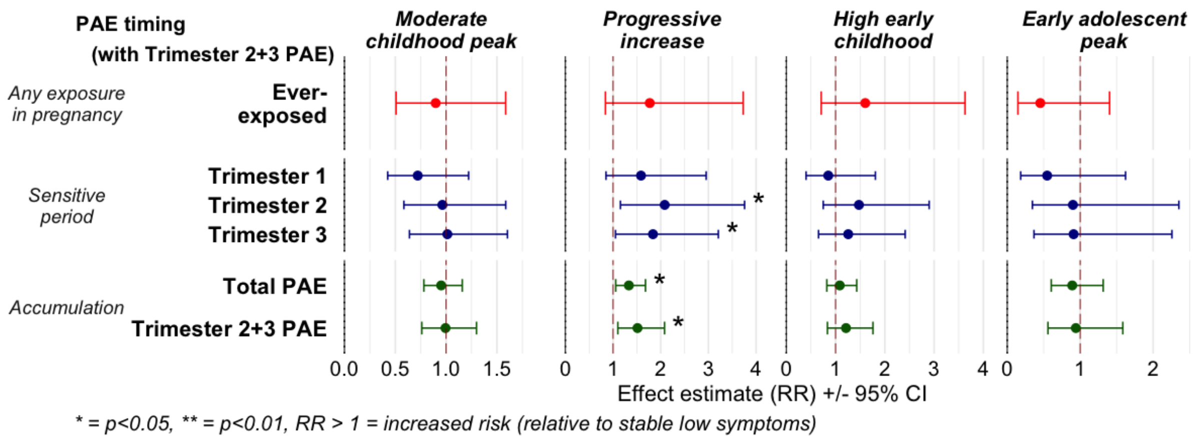

**Figure S4: Associations between main PAE and alternative accumulative PAE models with latent depressive symptom trajectories (relative risk ratios RR, 95% CI).** Risk conferred by accumulative burden in the second and third trimesters was compared to those by main PAE models. For sensitive periods, RR reflects risk conferred by each stepwise increase in PAE level (from none to low, or low to high). Top to bottom: any exposure in pregnancy, sensitive period exposure, accumulative burden across pregnancy or in second and third trimesters.

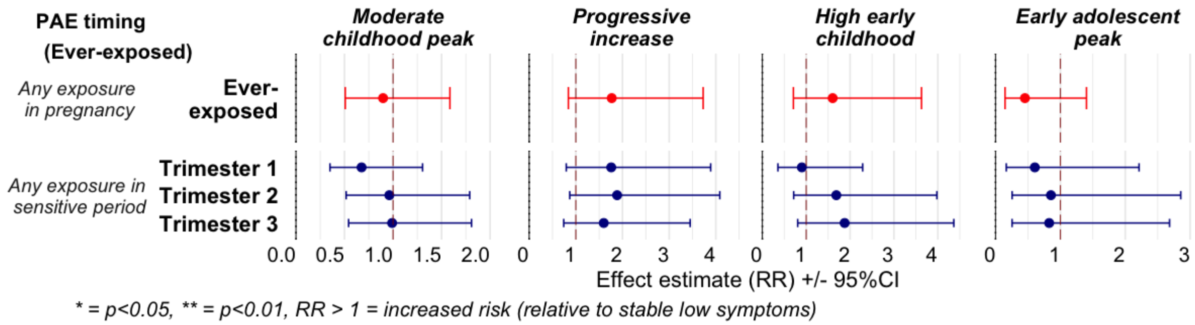

**Figure S5: Associations between ever-exposed PAE models and latent depressive symptom trajectories (relative risk ratios RR, 95% CI).** Dichotomous PAE models were used to evaluate if timing alone (i.e., without dosage information) was associated with depressive symptom trajectories. RR reflects risk conferred by any PAE during pregnancy or in each of the respective trimesters. Top to bottom: associations with any exposure in pregnancy, any exposure in sensitive periods.

**Appendix 6: Sensitivity analyses within trajectory construction sample (N = 6,409).**

We conducted regression analyses between PAE models and depressive symptom trajectories within the trajectory construction sample, which contained missingness in PAE reports. This analysis ensured that findings from the analytic sample (N=2,254) with complete trimester PAE reports were generalizable to a larger population, limiting concerns of potential selection bias.

**Table S6**: **Relative risk ratio (RR) conferred by PAE models for each depressive symptom trajectory compared to the stable low class.** 6,409 participants of the trajectory construction sample with incomplete PAE reports are included. N=number of participants with valid PAE measures.

| PAE models | Moderate childhood peak | Progressive increase | High early childhood | Early adolescent peak |
|---|---|---|---|---|
| | | *Timing hypotheses* | | |
| ***Ever-exposed*** | | | | |
| | 0.94 [0.67, 1.3]<br>p=0.70 | 0.95 [0.60, 1.48]<br>p=0.81 | 1.05 [0.64, 1.71]<br>p=0.85 | 0.55 [0.28, 1.08]<br>p=0.084 |
| ***Sensitive period*** | | | | |
| First trimester | 0.73 [0.49, 1.08]<br>p=0.12 | 1.05 [0.63, 1.75]<br>p=0.86 | 0.87 [0.49, 1.54]<br>p=0.64 | 0.72 [0.33, 1.54]<br>p=0.40 |
| Second trimester | 0.98 [0.73, 1.31]<br>p=0.87 | 1.25 [0.86, 1.83]<br>p=0.24 | 1.09 [0.71, 1.67]<br>p=0.70 | 0.71 [0.39, 1.29]<br>p=0.26 |
| Third trimester | 0.9 [0.64, 1.28]<br>p=0.57 | 1.28 [0.81, 2.02]<br>p=0.29 | 1.21 [0.74, 1.96]<br>p=0.44 | 0.76 [0.37, 1.55]<br>p=0.45 |
| ***Accumulation*** *(N=2,254)* | | | | |
| Pregnancy | 0.95 [0.78, 1.16]<br>p=0.62 | **1.33 [1.06, 1.68] *<br>p=0.016** | 1.08 [0.82, 1.43]<br>p=0.57 | 0.89 [0.60, 1.32]<br>p=0.56 |
| Second + third trimester | 0.99 [0.76,1.30]<br>p=0.97 | **1.51 [1.10, 2.08] *<br>p=0.011** | 1.21 [0.83, 1.76]<br>p=0.31 | 0.94 [0.56, 1.58]<br>p=0.82 |
| | | *Dosage hypotheses* | | |
| ***First trimester*** *(N=3,582)* | | | | |
| Low PAE | 0.93 [0.72, 1.20]<br>p=0.57 | 0.97 [0.68, 1.38]<br>p=0.86 | 0.96 [0.64, 1.42]<br>p=0.82 | 0.90 [0.54, 1.50]<br>p=0.68 |
| High PAE | 0.61 [0.35, 1.06]<br>p=0.08 | 1.14 [0.61, 2.16]<br>p=0.68 | 0.83 [0.40, 1.73]<br>p=0.62 | 0.64 [0.22, 1.83]<br>p=0.40 |
| ***Second trimester*** *(N=6,220)* | | | | |
| Low PAE | 1.04 [0.86, 1.26]<br>p=0.68 | 0.95 [0.73, 1.23]<br>p=0.68 | 1.0 [0.75, 1.34]<br>2.0 p=0.99 | 0.84 [0.57, 1.25]<br>p=0.39 |
| High PAE | 0.87 [0.58, 1.31]<br>p=0.52 | **1.61 [1.04, 2.51] *<br>p=0.034** | 1.18 [0.69, 2.02]<br>p=0.56 | 0.70 [0.30, 1.64]<br>p=0.41 |
| ***Third trimester*** *(N=3,940)* | | | | |
| Low PAE | 0.87 [0.68, 1.11]<br>p=0.26 | 0.93 [0.66, 1.30]<br>p=0.66 | 1.34 [0.96, 1.87]<br>p=0.08 | 0.99 [0.61, 1.59]<br>p=0.96 |
| High PAE | 1.02 [0.68, 1.54]<br>p=0.93 | 1.59 [0.97, 2.62]<br>p=0.07 | 0.88 [0.45, 1.72]<br>p=0.71 | 0.58 [0.21, 1.65]<br>p=0.31 |

RR; [95%CI]; p-value. RR: relative risk ratio, derived from exponentiated log odds ratio.

*p<0.05, **p<0.01.

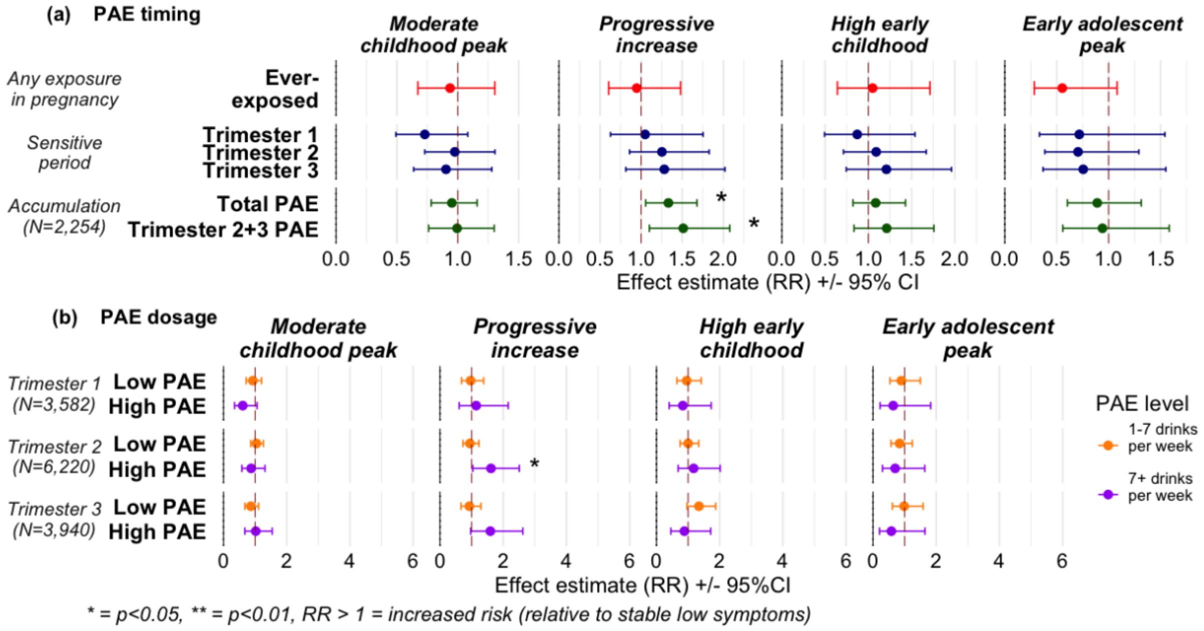

**Figure S6**: **Associations between PAE models and latent depressive symptom trajectories (relative risk ratios RR, 95%CI) within the trajectory construction sample.** 6,409 participants of the trajectory construction sample with incomplete PAE reports are included. N: number of participants with valid PAE measures. (**a)** Associations with PAE timing models (any exposure in pregnancy, sensitive period exposure, accumulative burden across pregnancy or in the second and third trimesters). For sensitive periods, RR reflects risk conferred by each stepwise increase in PAE level (from none to low, or low to high). (**b)** Associations with PAE dosage models (PAE at low and high dosages in the first, second and third trimesters of pregnancy).

**Appendix 7: Negative control analyses using partner drinking measures (N=6,409).**

As a negative control analysis, we tested for associations between timing (second/third trimester) and levels (no/low/high drinking) of partner drinking with depressive symptom trajectories, to confirm that risk differences in trajectory membership were contributed by PAE though maternal alcohol consumption.

**Table S7**: **Relative risk ratio (RR) conferred by partner drinking measures for each depressive symptom trajectory compared to the stable low class.** 6,409 participants of the trajectory construction sample with incomplete PAE reports are included. N: number of participants with valid partner drinking measures.

| **Partner drinking timing/level** | **Moderate childhood peak** | **Progressive increase** | **High early childhood** | **Early adolescent peak** |
|---|---|---|---|---|
| | | *Timing* | | |
| ***Sensitive period*** | | | | |
| Second trimester | 0.70 [0.47, 1.05] p=0.08 | 0.64 [0.36, 1.12] p=0.12 | 0.64 [0.35, 1.17] p=0.15 | 0.63 [0.28, 1.45] p=0.28 |
| Third trimester | 0.55 [0.18, 1.71] p=0.30 | 0.67 [0.15, 2.99] p=0.60 | 0.88 [0.19, 4.04] p=0.87 | 0.70 [0.10, 5.00] p=0.72 |
| | | *Drinking levels* | | |
| ***Second trimester*** *(N=4,950)* | | | | |
| Low drinking (N = 3739) | 0.73 [0.46, 1.15] p=0.18 | 0.99 [0.49, 1.99] p=0.98 | 0.76 [0.40, 1.44] p=0.40 | 0.67 [0.29, 1.51] p=0.33 |
| High drinking (N=1036) | 0.64 [0.39, 1.04] p=0.07 | 0.74 [0.35, 1.58] p=0.44 | 0.62 [0.30, 1.24] p=0.18 | 0.56 [0.23, 1.41] p=0.22 |
| ***Third trimester*** *(N=494)* | | | | |
| Low drinking (N = 338) | 0.76 [0.25, 2.28] p=0.62 | 0.32 [0.10, 1.03] p=0.06 | 0.82 [0.19, 3.43] p=0.78 | 0.47 [0.09, 2.49] p=0.37 |
| High drinking (N =118) | 0.55 [0.16, 1.90] p=0.34 | 0.45 [0.12, 1.72] p=0.25 | 0.81 [0.16, 4.14] p=0.80 | 0.54 [0.08, 3.55] p=0.52 |

RR; [95%CI]; p-value. RR: relative risk ratio, derived from exponentiated log odds ratio.

*p<0.05, **p<0.01.

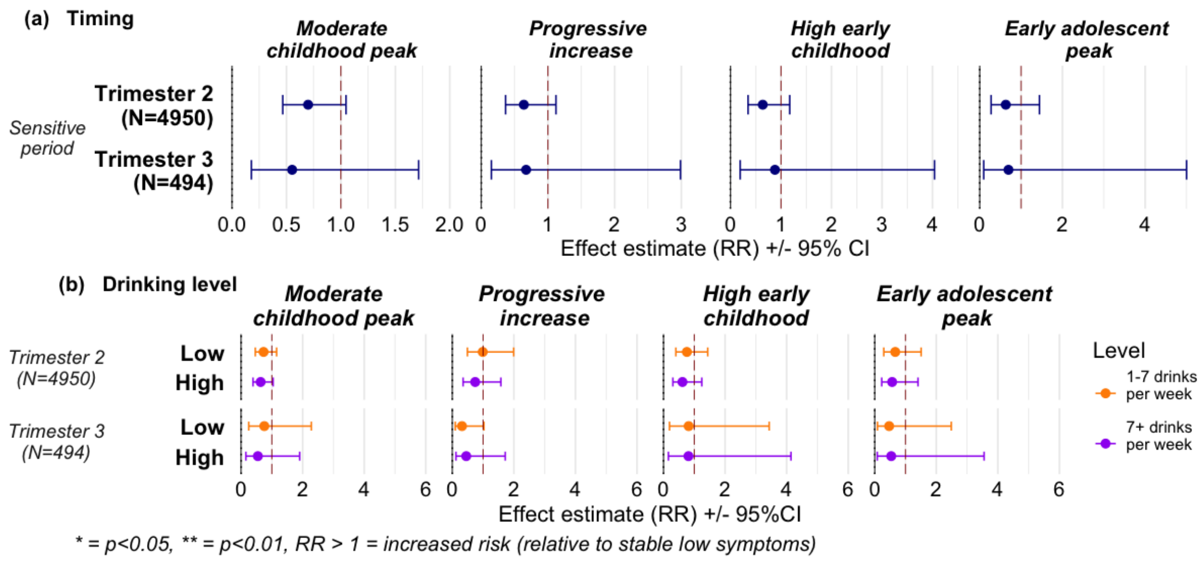

**Figure S7**: **Associations between partner drinking timing and levels with latent depressive symptom trajectories (relative risk ratios RR, 95% CI).** 6,409 participants of the trajectory construction sample are included. N: number of participants with valid partner drinking reports. There were insufficient partner drinking reports in the first trimester to investigate associations with latent depressive symptom trajectories. **(a)** Associations with partner drinking timing (sensitive period exposure in second or third trimester). For sensitive periods, RR reflects risk conferred by each stepwise increase in drinking level (from none to light, or light to heavy). **(b)** Associations with partner drinking levels (light or heavy drinking in the second or third trimester of pregnancy).

### Appendix 8: Participant demographics across PAE levels within the analytic sample (N=2,254).

**Table S8: Demographic characteristics for 2,254 analytic sample participants, stratified by overall PAE levels across pregnancy.** Total PAE burden across pregnancy was stratified into four levels: level 0 – never exposed in pregnancy; levels 1-3 – Low PAE in 1-2 trimesters; levels 4-5 – High PAE in 1-2 trimesters; level 6 – High PAE throughout pregnancy.

| | Never exposed in pregnancy (N=1,193) | Low PAE in 1-2 trimesters (N=910) | High PAE in 1-2 trimesters (N=116) | High PAE throughout pregnancy (N=35) | Overall (N=2,254) |
|---|---|---|---|---|---|
| **Gestational length (weeks)** | | | | | |
| Mean (SD) | 39.5 (1.57) | 39.6 (1.53) | 39.6 (1.63) | 39.9 (1.38) | 39.5 (1.55) |
| Median [Min, Max] | 40.0 [32.0, 43.0] | 40.0 [32.0, 44.0] | 40.0 [35.0, 44.0] | 40.0 [36.0, 42.0] | 40.0 [32.0, 44.0] |
| **Maternal age at birth** | | | | | |
| Mean (SD) | 28.9 (4.27) | 29.5 (4.27) | 30.1 (4.76) | 30.1 (4.89) | 29.2 (4.32) |
| Median [Min, Max] | 29.0 [16.0, 44.0] | 29.0 [17.0, 44.0] | 29.0 [21.0, 42.0] | 30.0 [21.0, 41.0] | 29.0 [16.0, 44.0] |
| **Child sex** | | | | | |
| Female | 588 (49.3%) | 451 (49.6%) | 57 (49.1%) | 21 (60.0%) | 1117 (49.6%) |
| Male | 605 (50.7%) | 459 (50.4%) | 59 (50.9%) | 14 (40.0%) | 1137 (50.4%) |
| **Parity** | | | | | |
| Mean (SD) | 0.742 (0.896) | 0.742 (0.835) | 0.776 (0.835) | 0.943 (0.873) | 0.747 (0.868) |
| Median [Min, Max] | 1.00 [0, 6.00] | 1.00 [0, 5.00] | 1.00 [0, 4.00] | 1.00 [0, 4.00] | 1.00 [0, 6.00] |
| **Ethnicity** | | | | | |
| White | 1157 (97.0%) | 884 (97.1%) | >111 (>95.7%) | >30 (>85.7%) | 2190 (97.2%) |
| Non-white | 36 (3.0%) | 26 (2.9%) | <5 (<4.3%) | <5 (<14.3%) | 64 (2.8%) |
| **Maternal education at birth** | | | | | |
| Below O-level | 234 (19.6%) | 136 (14.9%) | 21 (18.1%) | 10 (28.6%) | 401 (17.8%) |
| O-level | 467 (39.1%) | 337 (37.0%) | 41 (35.3%) | 14 (40.0%) | 859 (38.1%) |
| A-level | 328 (27.5%) | 250 (27.5%) | 28 (24.1%) | 6 (17.1%) | 612 (27.2%) |
| Tertiary degree or above | 164 (13.7%) | 187 (20.5%) | 26 (22.4%) | 5 (14.3%) | 382 (16.9%) |
| **EPDS 8 weeks postpartum** | | | | | |
| Mean (SD) | 5.38 (4.68) | 5.54 (4.27) | 6.76 (5.01) | 8.14 (6.47) | 5.56 (4.59) |
| Median [Min, Max] | 4.00 [0, 28.0] | 5.00 [0, 27.0] | 6.00 [0, 23.0] | 7.00 [0, 25.0] | 5.00 [0, 28.0] |
| **Smoking during pregnancy** | | | | | |
| Yes | 175 (14.7%) | 134 (14.7%) | 22 (19.0%) | 13 (37.1%) | 344 (15.3%) |
| No | 1018 (85.3%) | 776 (85.3%) | 94 (81.0%) | 22 (62.9%) | 1910 (84.7%) |

**Appendix 9: Average modal probabilities of trajectory class predictions.**

Using the most probable class membership from the GMM as the outcome of multinomial logistic regressions disregards any class prediction uncertainty, which adds to measurement error. To characterize the extent of uncertainty in the class predictions, we collect the exact class prediction probabilities for all participants, as well as their final assignment, and calculate the average prediction probability for each of the five latent classes. The higher the certainty of the class predictions, the closer the average will be to 1.

We computed and compared the statistic for the trajectory construction sample versus analytic sample with complete PAE reports used in regression analyses.

**Table S9: Average modal probabilities of trajectory class predictions in the trajectory construction sample and analytic sample.**

| Trajectory class | Average modal probabilities for trajectory sample (N = 6,409) | Average modal probabilities for analytic sample (complete PAE; N = 2,254) |
|---|---|---|
| Stable low | 0.939 | 0.942 |
| High early childhood | 0.753 | 0.795 |
| Early adolescent peak | 0.904 | 0.923 |
| Progressive increase | 0.789 | 0.798 |
| Moderate childhood peak | 0.790 | 0.779 |
